# Final-size solutions for SIRI models with vaccination

**DOI:** 10.1101/2025.09.12.25335631

**Authors:** Maria A. Gutierrez, Julia R. Gog

## Abstract

In the classic SIR model, infection gives full immunity against any possible reinfection. However, for many important epidemiological situations, immunity is only partial and reinfection is possible. Though these models are mathematically more complex, we are able to find expressions for the epidemic final size. We also generalise these expressions to include vaccination, with a fraction of the population vaccinated before the epidemic, where vaccinees are less susceptible to primary infections than unvaccinated hosts.

Partial immunity can be interpreted at the population level as providing either full or no protection to each host, in some proportion (all-or-none immunity). In this scenario, we give analytical expressions (mathematically similar to the SIR final-size) for the cumulative primary infections and the cumulative reinfections in unvaccinated and vaccinated hosts. Alternatively, partial immunity can be interpreted as providing homogeneous imperfect protection to each host (leaky immunity). For this other scenario, we again obtain an implicit equation for the final epidemic size. We break down, in terms of the final size, the number of infections in hosts with or without prior immunity (vaccine- or infection-induced), as well as the number of primary infections and reinfections. Under the leaky immunity assumption, we find a form of reinfection threshold. If the relative host susceptibility to reinfection is above this threshold (which is the inverse of the pathogen’s basic reproduction number), transmission rates are high enough to support an endemic disease. Below the reinfection threshold, epidemics are transient. In the all-or-none model, epidemics are always transient.

## 1 Introduction

The classic SIR epidemic model, first analysed in [32], includes the simplifying assumption that infection confers total immunity [20]. However, many pathogens do not provide complete immunity: reinfection is possible [8]. Moreover, there is some evidence of sequential infections with the same strain in both Influenza A [26] and SARS-CoV-2 [34], although waning immunity may play an important role here. The Omicron SARS-CoV-2 variant provides especially low protection against reinfection [30]. Reinfections can be key in shaping the epidemic dynamics of some pathogens, such as SARS-CoV-2 [22]. As a rule of thumb, epidemics will be prolonged if reinfection is possible. Furthermore, some pathogens become endemic due to the lack or loss of protection against reinfection [14].

Deterministic compartmental models are widely used in epidemiology to study the spread of infectious diseases [20]. A key advantage of these simple ODE models is that they allow direct access to the “final size” of an epidemic: the cumulative number of infections in the host population. Final-size expressions allow more complex phenomena to be explored; for example, the evolution of a disease across multiple seasons [1, 3] or the evolutionary pressure acting on a pathogen over a single epidemic wave [17]. An expression for the final size of a classical SIR epidemic can be derived in terms of the Lambert W function [24], and this final-size solution can be generalised in various ways [10, 17, 25].

A common relaxation of the full immunity assumption is that infection induces full but temporary immunity [20]. The biological mechanism here could be loss of immunity, or the antigenic evolution of the circulating pathogen [2, 29]. These SIRS-type models do not usually have final-size solutions because the epidemic does not burn out. Instead, a positive endemic equilibrium is possible because waning of immunity replenishes the pool of susceptible individuals, offering a dynamic balance to susceptible depletion through infection [20].

In this paper, we instead consider the alternative assumption of lifelong but imperfect immunity: infection offers some protection against further infections, but reinfection is still possible. These “SIRI”-type epidemic dynamics often show a transient epidemic. We obtain analytic final-size expressions for the total infections over the course of the epidemic, split into primary infections and reinfections. This is also extended to vaccination, and then the final size is further disaggregated by host vaccination status. We consider two interpretations of the “partial” nature of immunity: “all-or-none” and “leaky” immunity [13].

## 2 General modelling assumptions

We use deterministic compartmental models to find the cumulative number of infections during an epidemic wave. We assume a single strain of an infectious disease and a well-mixed population of constant size, with no births or deaths. We choose time units to make the recovery rate one, which we assume is the same for all hosts (regardless of immune status). We consider an epidemic that is unmitigated, apart from the use of vaccines. We assume that a proportion *c* of the population is vaccinated before the epidemic. We consider only vaccines that provide partial but lifelong immunity (explained below), so vaccinated individuals may still become infected. Up to this point, these assumptions follow those in [17].

We assume that both infection- and vaccine-induced immunity reduce host susceptibility but not necessarily by the same amount, similarly to the “partial immune protection” model of [14]. We use *θ*_*S*_ to denote the relative susceptibility of vaccinees: *θ*_*S*_ = 1 means no protection against infection. Analogously, we use *ϕ*_*S*_ for the relative susceptibility of recovered hosts (*ϕ*_*S*_ = 1 means no protection against reinfection). Thus, unless *ϕ*_*S*_ = 0, reinfections are possible, in contrast to [17] where recovered individuals are fully protected. We assume that prior immunity does not change host infectiousness.

**Table 1.** Parameters and output functions.

| item | description |
| --- | --- |
| $\theta_S \in [0, 1]$ | relative susceptibility (to a first infection) of vaccinees |
| $\phi_S \in [0, 1]$ | relative susceptibility of recovered hosts, analogous to $\theta_S$ |
| $c \in [0, 1]$ | vaccination coverage of the population |
| $R_0$ | initial basic reproduction number, assumed $R_0 > 1$ |
| $R_e(c)$ | $R_e = R_0(1 - c(1 - \theta_S))$, initial effective reproduction number, assumed $R_e > 1$ |
| $c_h$ | vaccination coverage threshold for $R_e(c_h) = 1$, $c_h = (1 - R_0^{-1})/(1 - \theta_S)$ |
| $r(c)$ | $(R_e - \phi_S)/(1 - \phi_S)$, similar to $R_e$, but including reinfections for $\phi_S > 0$ |
| $C_U(c)$ | total primary infections in unvaccinated hosts |
| $C_V(c)$ | total primary infections in vaccinated hosts |
| $C_U^\dagger(c)$ | total reinfections in unvaccinated hosts |
| $C_V^\dagger(c)$ | total reinfections in vaccinated hosts |

## 3 Model with all-or-none immunity

### 3.1 Model description

For the all-or-none model assumption, the relative susceptibility factors (*θ*_*S*_ and *ϕ*_*S*_) are interpreted as all-or-none immunity (also termed “polarised” or “all-or-nothing” immunity) [11, 13, 18]. With all-or-none vaccine-induced immunity, a proportion 1 − *θ*_*S*_ of the vaccinated individuals become fully protected against the disease. The remaining *θ*_*S*_ vaccinated individuals are still fully susceptible to the disease. Similarly, after infection, an individual becomes fully immune with probability 1 − *ϕ*_*S*_ or returns to the susceptible compartment with probability *ϕ*_*S*_. Section 4 instead uses leaky immunity, as in [14] and other “SIRI”-type (Susceptible-Infected-Recovered-Infected) models (e.g., [28]).

We refer to the vaccinated and unvaccinated with the subscripts *U* and *V* . We use the superscript † for individuals who have been infected before. The compartments of the model are thus, 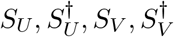, the proportions of the population fully susceptible to the disease, and *I*_*U*_, *I*_*V*_, the infected proportions. *R*_0_ is the basic reproduction number and *c* is the vaccinated proportion of the population. The ODE model is

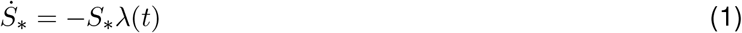

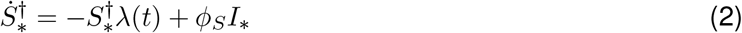

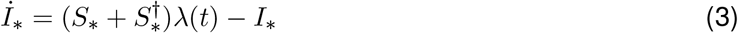

for each of * = *U, V* and *λ*(*t*) = *R*_0_(*I*_*U*_ +*I*_*V*_ ) is the force of infection and the dots denote differentiation with respect to time. We assume that all unvaccinated individuals are initially susceptible, except for a few infections that initiate the outbreak. Similarly, we assume that a proportion *θ*_*S*_ of those vaccinated is fully susceptible from the start of the outbreak (as in the all-or-none models of [17]). Therefore, the initial conditions are

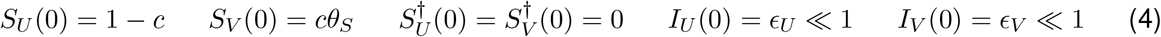

where *ϵ*_*U*_, *ϵ*_*V*_ ≥ 0 represent the infected hosts that initiate the outbreak (at least one of *ϵ*_*U*_, *ϵ*_*V*_ is positive). The final-size calculation (Section 3.2) later sets these infinitesimal proportions of the population to zero, as in the standard approach to final-size calculations [20].

### 3.2 Final-size calculation

The vaccinated and unvaccinated populations are subject to the same epidemic dynamics (1)-(3). Thus, they stay locked in proportion to each other (as per Appendix A.1 of [17]), according to the initial conditions (4):

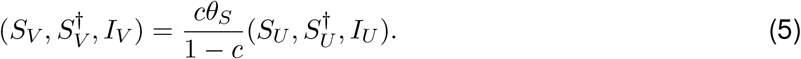

provided that (1 − *c*)*ϵ*_*V*_ = *cθ*_*S*_*ϵ*_*U*_ (which we assume to be true since *ϵ*_*U*_, *ϵ*_*V*_ ≪ 1). We define new variables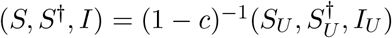 and use (5) to obtain

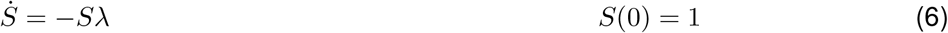

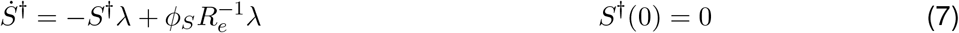

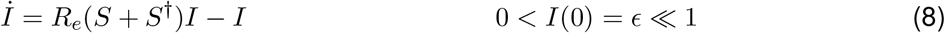

where *λ* = *R*_*e*_*I* and *R*_*e*_ = *R*_0_[1 − *c*(1 − *θ*_*S*_)] is the initial effective reproduction number. For *θ*_*S*_ < 1, increasing the vaccinatoon coverage *c* lowers *R*_*e*_. If the vaccination coverage is above its critical value 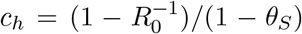, the effective reproduction number is less than one (*R*_*e*_ < 1). Therefore, herd immunity prevents an outbreak for a vaccination coverage above this threshold (*c* > *c*_*h*_): the prevalence decreases exponentially. If, instead, the vaccination coverage is below the threshold (*c* < *c*_*h*_), the epidemic grows. To obtain the final size of the epidemic, we aim to express *I* in terms of *S* only (we use mathematical manipulations similar to those involved in the traditional derivation of the SIR final-size result, as the models here are not amenable to analysis with some other final-size approaches [27]). Using the chain rule,

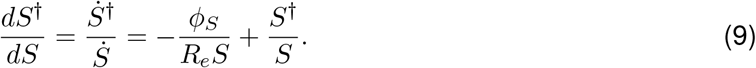

Multiplying both sides of (9) by 1*/S* we see that 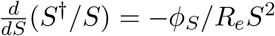, which we integrate using the initial conditions *S*^†^ = 0 and *S* = 1 at *t* = 0. Hence, *S*^†^*/S* = *ϕ*_*S*_(1*/S* − 1)*/R*_*e*_ and thus

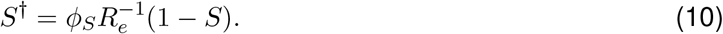

As expected, *S*^†^ in (10) is a decreasing function of *S*, because *S* (a proxy for the proportion of hosts who have never been infected) continuously decreases throughout the epidemic while *S*^†^ (recovered hosts) increases. Using (10), the ODE for *I*(*t*) in (8) becomes

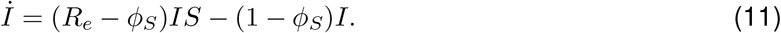

By comparison with the standard SIR equations, the ODE (11) suggests that *r* = (*R*_*e*_ − *ϕ*_*S*_)*/*(1 − *ϕ*_*S*_) may determine the epidemic dynamics here in a similar way to how the basic reproduction number *R*_0_ = *β/γ* affects the SIR dynamics: Section 3.3 discusses this analogy further. To calculate the final epidemic size, we use (11) and (6):

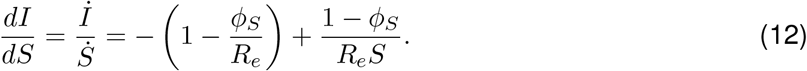

Using the initial conditions *I* = 0 and *S* = 1 at *t* = 0, we obtain

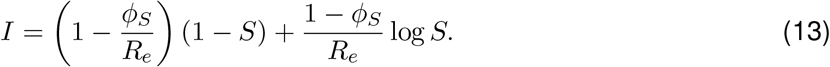

To find the final size *S*^∞^ := lim_*t*→∞_ *S*(*t*) < 1, we take *t* → ∞, and set lim_*t*→∞_ *I*(*t*) = 0. Thus,

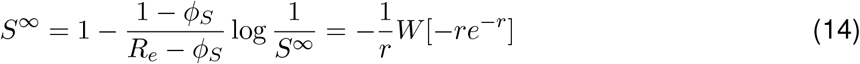

where *r* = (*R*_*e*_ − *ϕ*_*S*_)*/*(1 − *ϕ*_*S*_), as above, and *W* is the (principal branch of the) Lambert W function [24]: obeying *W*(*x*)*e*^*W*(*x*)^ and *W* (*x*) ∈ [−1, 0] for *x* ∈ [*e*^−1^, 0]. The cumulative number of primary infections in the unvaccinated *C*_*U*_ = (1 − *c* − lim_*t*→∞_ *S*_*U*_). Since *C*_*U*_ counts infections across the full epidemic, it does not depend on the time *t*. By definition *S*_*U*_ = (1−*c*)*S*, hence *C*_*U*_ = (1−*c*)(1−*S*^∞^) = (1 − *c*) (1 + *W*[−*re*^−*r*^]*/r*). The cumulative reinfections in the unvaccinated are

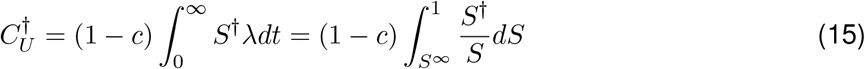

which we integrate using (10) and (14), so that 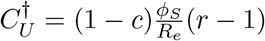 (1 + *W*[−*re*^−*r*^]*/r*).

### 3.3 Final-size results

For vaccination coverages below the herd-immunity threshold (*c* < *c*_*h*_), the calculations above show that the cumulative primary infections in unvaccinated individuals are

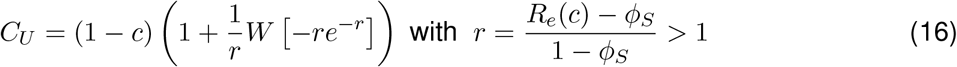

where *W* is the Lambert W function [24] and 0 ≤ *ϕ*_*S*_ < 1. The proportions (5) imply that the cumulative primary infections in vaccinated individuals are *C*_*V*_ = *cθ*_*S*_*C*_*U*_*/*(1 − *c*). If past infections do not protect at all against future infections (*ϕ*_*S*_ = 1), the final-size calculation (16) is not valid (because *r* = ∞). Instead, with *ϕ*_*S*_ = 1, we have an SIS model [20]. For *r* ∈ (1, ∞), the factor *f* (*r*) = (1 + *W* [− *re*^−*r*^] */r*) in (16) is the final size of a standard SIR epidemic with initial reproduction number *r* [24]. Therefore, even though *R*_*e*_ is the actual initial effective reproduction number, *r* plays a more important role here with reinfections (*ϕ*_*S*_ > 0), as the ODE (11) suggests. Unfortunately, we do not have a clear biological interpretation of *r*. Without reinfections (*ϕ*_*S*_ = 0), *r* is just *R*_*e*_.

As the probability of remaining susceptible upon recovery (*ϕ*_*S*_) increases from zero, transmission increases: *r* > *R*_*e*_ monotonically increases with increasing *ϕ*_*S*_. Therefore, with reinfections (*ϕ*_*S*_ > 0) there are more primary infections *C*_*U*_, *C*_*V*_ than without reinfections (*ϕ*_*S*_ = 0). Both *C*_*U*_ and *C*_*V*_ monotonically increase with increasing *ϕ*_*S*_, because a higher *ϕ*_*S*_ requires more infections in the population to build herd immunity (which overturns the epidemic).

We have also shown above that the cumulative reinfections in unvaccinated hosts are

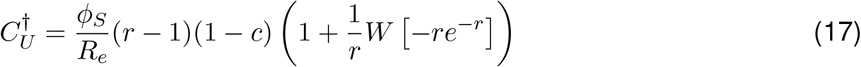

for *c* < *c*_*h*_. Using (5), 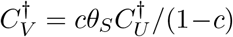 are the reinfections in vaccinated individuals. Unsurprisingly both 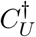 and 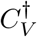 are increasing functions of *ϕ*_*S*_ (lower protection against reinfection means there are more reinfections).

The final-size expressions (16) and (17) show that the ratio of reinfections to primary infections is

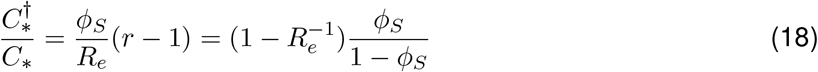

for both the unvaccinated (∗ = *U* ) and vaccinated (∗ = *V* ) subpopulations. This ratio (18) can take any positive value as *ϕ*_*S*_ varies from zero (full protection against reinfection) to one (no protection). In particular, if the protection against reinfection is low (*ϕ*_*S*_ ≈ 1), there may be more reinfections than primary infections. Interestingly, this ratio of secondary to primary infections decreases as the vaccination coverage *c* increases. In other words, with more vaccinated individuals, a higher proportion of the total infections are primary infections. The disease has fewer opportunities to reinfect hosts, because a higher vaccination coverage means that the final epidemic size is smaller.

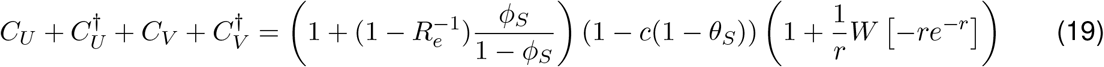

are the total infections. In the limit *R*_0_ → ∞, the right-most factor above tends to 1 (meaning that in the classic SIR model, the full population becomes infected), and so (19) tends to (1 − *c*(1 − *θ*_*S*_))/(1 −*ϕ*_*S*_). This limit can also be derived from first principles: 1 −*c*(1 −*θ*_*S*_) is the initial susceptible size and 1/(1 −*ϕ*_*S*_) = 1 +*ϕ*_*S*_(1 +*ϕ*_*S*_(… )) is the expected number of infections an individual undergoes before becoming immune (this number of infections follows a geometric distribution).

#### Final-size results without vaccination

Setting *c* = 0 gives the final-size results in the absence of vaccination. Assuming *R*_0_ > 1, the cumulative primary infections are

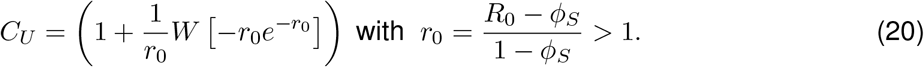

Similarly, the total reinfections are

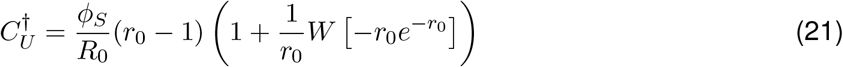

and the total infections are

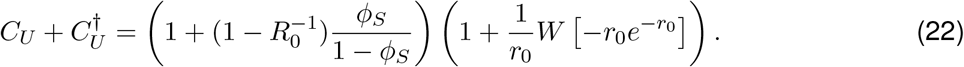

## 4 Model with leaky immunity

### 4.1 Model description

We modify the all-or-none model above to instead account for “leaky” immunity. In this scenario, both infection- and vaccine-induced immunity partially protect against infection, reducing individual host susceptibility but not fully preventing future infections. All hosts with partial immunity can become infected, though at lower rates than those with no immunity. Vaccinees who have never been infected have relative susceptibility *θ*_*S*_ ∈ [0, 1]. Recovered hosts have relative susceptibility *ϕ*_*S*_ ∈ [0, 1], regardless of vaccination status: we make this simplifying assumption for tractability. Biologically, we expect *ϕ*_*S*_ ≤ *θ*_*S*_ so that infection does not increase the susceptibility of vaccinated hosts. With these assumptions, the model becomes

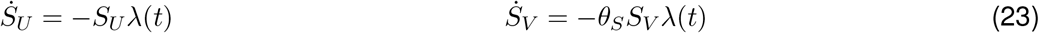

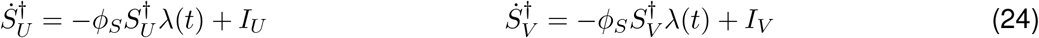

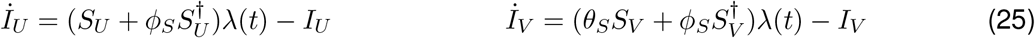

where *λ* = *R*_0_(*I*_*U*_ + *I*_*V*_ ) is the force of infection (as before). Initially, all vaccinees are susceptible to infection. Hence, 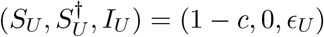 and 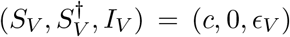 at *t* = 0 (again with 0 ≤ *ϵ*_*U*_, *ϵ*_*V*_ ≪ 1 and *ϵ*_*U*_ + *ϵ*_*V*_ > 0). The initial effective reproduction number is *R*_*e*_ = *R*_0_(1 − *c*(1 − *θ*_*S*_)), as before. Hence, the epidemic outbreak again grows for vaccination coverages *c* below the herd-immunity threshold 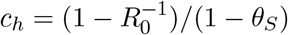.

### 4.2 Final-size equation

To derive the final size of the epidemic, the force of infection *λ* is expressed in terms of the number of susceptibles. Substituting for 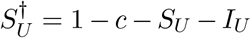 and 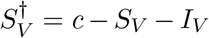 in 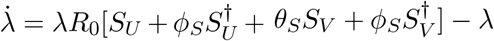:

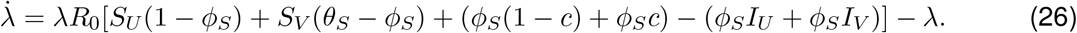

We want to express 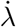 purely in terms of *λ* and *S*_*U*_ to obtain an implicit final-size equation. We eliminate *S*_*V*_ from (26) using

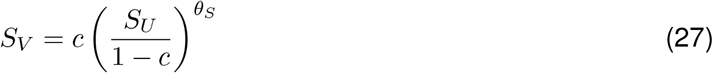

which follows from (23) and the initial conditions above. The relation (27) is as in Appendix A.2 of [17]. Substituting (27) in (26):

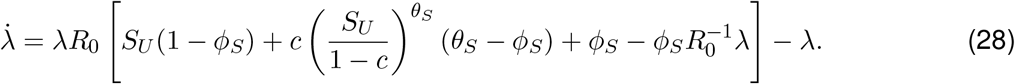

Using the chain rule and (28),

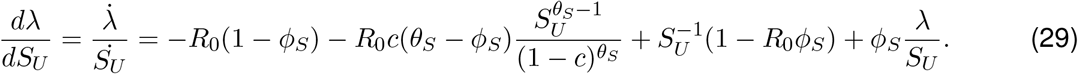

We solve (29) to find *λ* as a function of *S*_*U*_ :

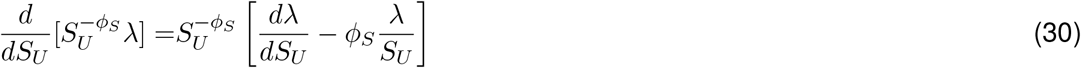

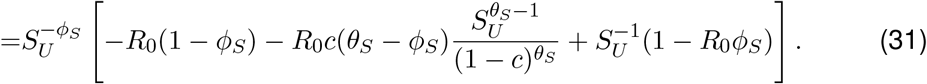

Therefore,

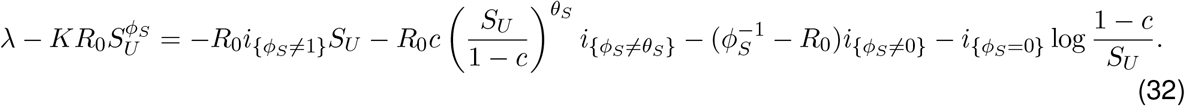

where *i*_{*Y* }_ is the indicator function (equal to one if the statement *Y* is true, and zero otherwise) and *K* is a constant of integration, set by the initial conditions *λ*(0) = 0 and *S*_*U*_ (0) = 1 − *c*:

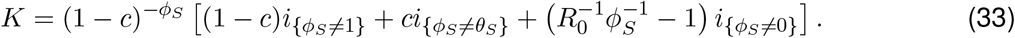

Therefore,

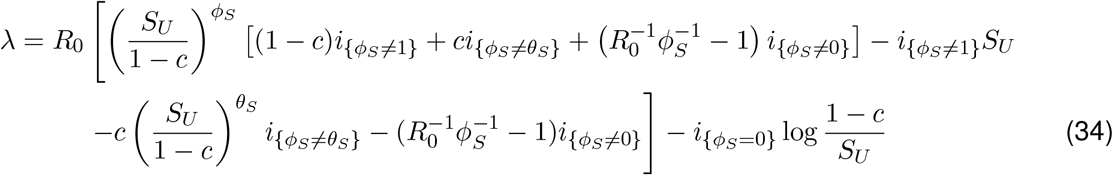

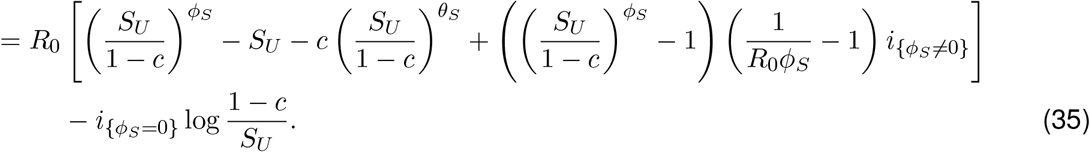

Setting lim_*t*→∞_ *λ*(*t*) = 0 in (35) gives an implicit equation for the final size 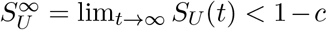. If *ϕ*_*S*_ = 0, reinfections are impossible. Thus, with *ϕ*_*S*_ = 0 we would obtain the final-size equation (43) of [17], here with *θ*_*I*_ = 1. For *ϕ*_*S*_ > 0, the final-size equation is

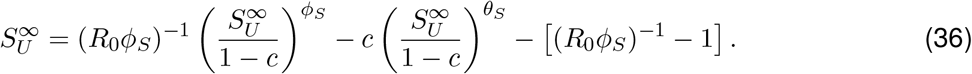

We show below that (36) has a solution 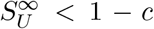 for *R*_0_*ϕ*_*S*_ < 1. If *R*_0_*ϕ*_*S*_ > 1, the epidemic reaches a stable endemic equilibrium—so *R*_0_ = 1*/ϕ*_*S*_ is a “reinfection threshold” [14, 15]—in which all hosts have been infected before (*S*_*U*_ = 0 = *S*_*V*_, analogous to the endemic equilibrium of the standard SIS model [20] for 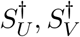 and *I*_*U*_, *I*_*V*_ ). In contrast, the all-or-none model does not have an endemic equilibrium (the last paragraph of Section 5 discusses this important difference). Here we are interested in a transient epidemic with both primary infections and reinfections, so henceforth we assume *R*_0_*ϕ*_*S*_ < 1.

#### Existence and uniqueness of the solution to the final-size equation

The initial condition *S*_*U*_ (0) = 1 − *c* for the susceptible size before the outbreak solves (36). We now prove that (36) also has a solution 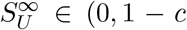, provided that *R*_0_ < 1*/ϕ*_*S*_ (see previous paragraph) and that the vaccination coverage is below the threshold for herd immunity 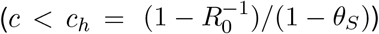. We define 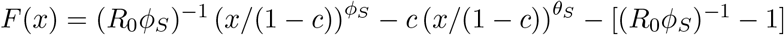 so that 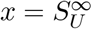 is a fixed point of *F* (*x*) = *x*. We also define *g*(*x*) = *F* (*x*) − *x*, so that *g*(1 − *c*) = 0 and

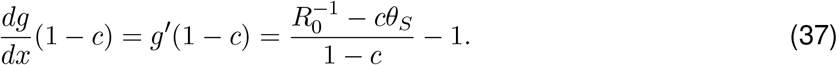

Using 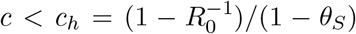, it follows that *g*^*′*^(1 − *c*) < 0. Since *g*(1 − *c*) = 0, we can pick *L* > 0 such that *g*(1 − *c* − *η*) > 0 for all *η* ∈ (0, *L*]. Moreover, *g*(0) = −[(*R*_0_*ϕ*_*S*_)^−1^ − 1] < 0. By the intermediate value theorem (applied to *g*(*x*), which is a continuous function of *x*), there exists a solution 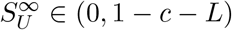 to 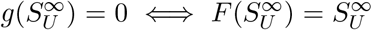, the final-size equation (36). To prove the uniqueness of 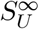, we use the mean value theorem. If *x*_1_ = *F*(*x*_1_) < *x*_2_ = *F*(*x*_2_), then there exists *x*_∗_ ∈ (*x*_1_, *x*_2_) such that *F*^*′*^(*x*_∗_) = (*F*(*x*_1_) − *F*(*x*_2_))*/*(*x*_1_ − *x*_2_) = 1. For *x*_∗_ > 0, *F*^*′*^(*x*_∗_) = 1 is equivalent to

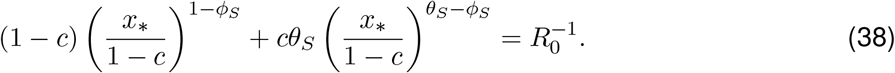

Since 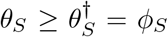, both exponents in (38) are non-negative. Therefore, the left-hand side of (38) monotonically increases with increasing *x*_∗_: from zero at *x*_∗_ = 0 (or if *θ*_*S*_ = *ϕ*_*S*_, from *cϕ*_*S*_ ≤ *ϕ*_*S*_ < 1*/R*_0_), to 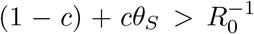 at *x*_∗_ = 1 − *c*. Hence, (38) has only one solution *x*_∗_ ∈ (0, 1 − *c*). Thus, *F* (*x*) has at most two fixed points *x*_1_, *x*_2_ ∈ [0, 1 − *c*]. Since 1 − *c* is a fixed point, we must have *x*_2_ = 1 − *c*, so 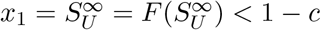 is unique.

### 4.3 Final-size results

For *R*_0_*ϕ*_*S*_ < 1, the final number of unvaccinated individuals who never become infected is the solution 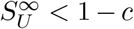 of (36). The cumulative primary infections are 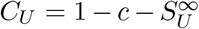 (in the unvaccinated) and 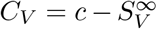 (in the vaccinated), with 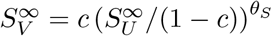 following from (27). Therefore, the total primary infections are

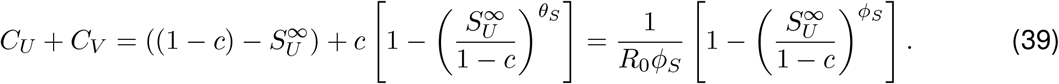

The total infections are

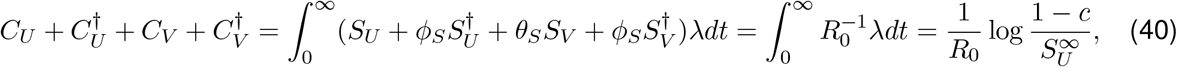

where we have used the expressions for 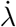 (given above (26)) and 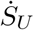. Thus, the cumulative infections in hosts with prior immunity are

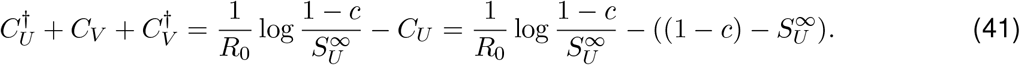

and—subtracting (39) from (40)—the cumulative reinfections are

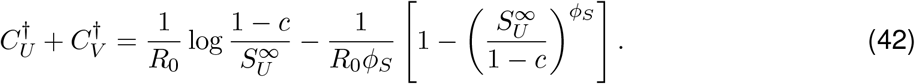

We have not been able to find expressions for 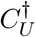 and 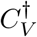 alone.

#### Final-size results without vaccination

Setting *c* = 0 simplifies the final-size equation (36), which is valid for *R*_0_*ϕ*_*S*_ < 1, to

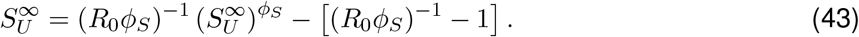

Thus, the cumulative primary infections are 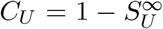, where 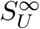 is the solution 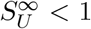 of (43). Similarly, the total infections are

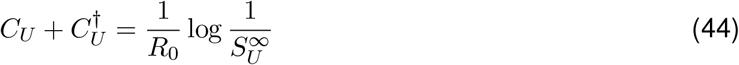

and the total reinfections are

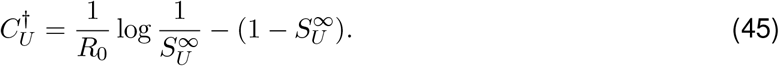

## 5 Discussion

In Section 3, we have studied an epidemic model including reinfections, in which immunity (from both infection and vaccination) is “all-or-none” [18]. The all-or-none immunity assumption facilitates the mathematical analysis, because there are inherently still only two immunity statuses in the model (fully immune or fully susceptible). While this form of immunity has been used to consider partial cross-immunity between distinct antigenic strains [9, 23], we are not aware of other models purely of reinfection that consider this all-or-none form.

The assumption that immunity against reinfection is “leaky” has been considered in other models before [12, 14, 15, 28]. Since we include vaccination, the model in Section 4 is most similar to the model for “partial immune protection and vaccination” of [14], but there are two key differences. First, because we do not model the birth-and-death dynamics of the population, we do not consider vaccination at birth. Instead, we capture the vaccination coverage of the population through the initial conditions of an epidemic (as in Section 3). Second, we assume the same susceptibility to reinfection for all hosts, regardless of vaccination status. We assume that this susceptibility to reinfection is not higher than the susceptibility of vaccinees who have never been infected. In contrast, [14] assumes that vaccinees always have the same susceptibility, regardless of whether they have been infected before or not, and that this susceptibility is lower than that of recovered uninfected individuals. Thus considers vaccines that protect against infections at least as much as infection-induced immunity, while Section 4 considers vaccines that protect no more than infection-induced immunity.

As in [14], we find a “reinfection threshold” (RT) for *R*_0_, separating two qualitatively different epidemiological scenarios. However, the RT in Section 4 is always determined by the relative susceptibility to reinfection *ϕ*_*S*_: unlike the RT of [14], the RT here does not depend on the relative susceptibility of vaccinees. Moreover, in [14], the RT only separates two quantitative regimes for the same endemic equilibrium, so it is not a bifurcation point of the dynamical system [4]. In contrast, the RT here is a bifurcation point as it separates a transient epidemic wave (*R*_0_ < 1*/ϕ*_*S*_) from an endemic equilibrium (*R*_0_ > 1*/ϕ*_*S*_). The endemic state occurs if the infection-induced protection against reinfection is low relative to the transmission rate 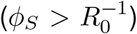. In this endemic equilibrium state, all hosts have previously been infected. Since this endemic state does not arise in the all-or-none model, this work highlights the importance of the mode in which immunity acts (leaky or all-or-none, although reality is likely somewhere between these two extremes [13]).

In both the all-or-none and leaky, a recovered host has the same relative probability (*θ*_*S*_) of becoming infected the first time it is re-challenged with infection. However, in subsequent challenges, the models differ. On the one hand, a population with leaky immunity always has the potential to be infected further under sufficient force of infection (each individual host, upon sufficient exposure to the pathogen, could be infected infinitely many times). On the other hand, the all-or-none form means a limit to the capacity for reinfection, with eventually all hosts fully immune under a sustained force of infection. This dynamic difference is analogous to the effects of partial cross-immunity between pathogen strains: sustained oscillations are possible with the leaky form under certain parameters, but never with the all-or-none form [7]. The parallel in this work is that the epidemic is always transient under reinfection with all-or-none immunity but can be sustained for leaky immunity. Similarly, paramaters leading to a transient epidemic under leaky immunity produce a smaller final epidemic size under all-or-none immunity (Figure 3).

**Figure 1.**
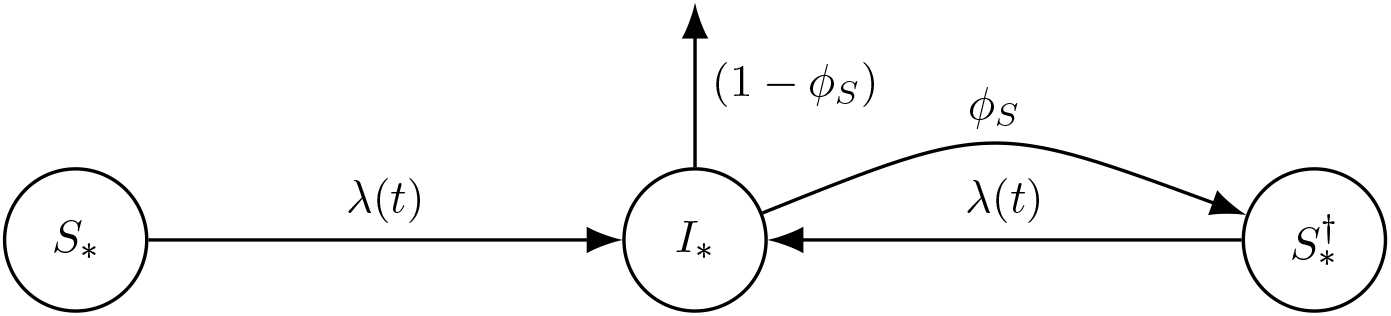
Schematic representation of the SIRI all-or-none model. The movement of individuals between compartments is analogous for the unvaccinated (∗ = *U*) and vaccinated (∗ = *V* ). Flow rates between compartments are given per capita (and with the average infectious period as time unit). The arrow with rate 1 − *ϕ*_*S*_ out of *I*_∗_ represents those hosts who become fully immune upon recovery from infection.

**Figure 2.**
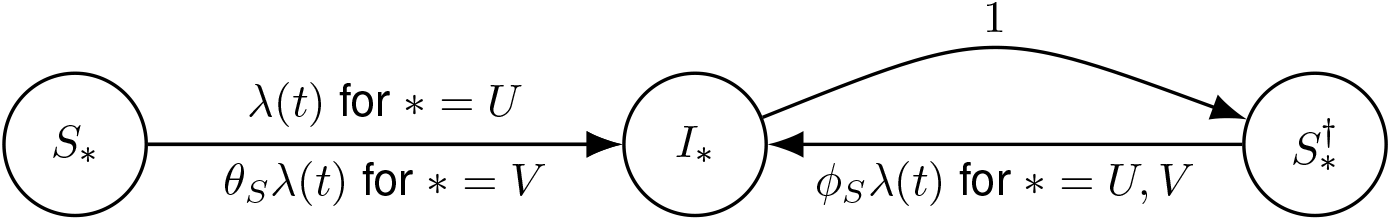
Schematic representation of the SIRI leaky model. The movement of individuals from *S*_∗_ to *I*_∗_ differs between the unvaccinated (∗ = *U*) and vaccinated (∗ = *V*), as vacinees have lower susceptibility. Flow rates between compartments are given per capita (and with the average infectious period as time unit).

**Figure 3.**
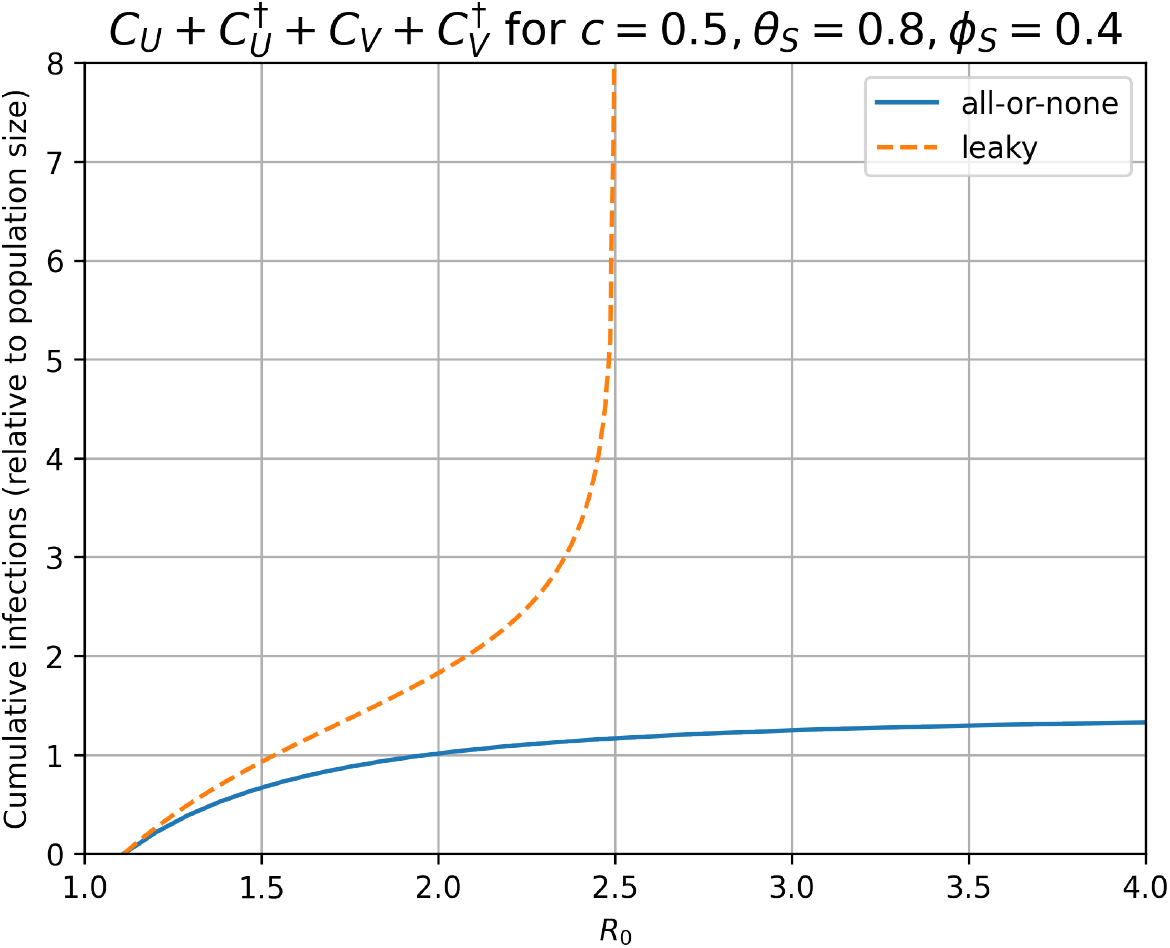
Cumulative number of infections for the leaky and all-or-none models, for fixed vaccination coverage *c* = 0.5, and susceptibilities *θ*_*S*_ = 0.8, *ϕ*_*S*_ = 0.4. We show values of *R*_0_ above 1/(1 −*c*(1 −*θ*_*S*_)) = 10/9 so that the initial effective reproduction number is greater than one (*R*_*e*_ > 1). With leaky immunity, the final size approaches infinity as *R*_0_ approaches the reinfection threshold (here 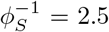). With all-or-none immunity, as *R*_0_ → ∞, the final size (19) approaches a constant (1 −*c*(1 −*θ*_*S*_))/(1 − *ϕ*_*S*_) = 3/2, as explained in Section 3.3.

Both epidemic models above group together all infectious hosts with the same vaccination status, regardless of how many times (if any) they have been infected before. This assumption is needed to limit the number of model compartments. Some of the implicit assumptions are that there are no differences between (i) host infectiousness during primary infections and reinfections, (ii) the infectious period during primary infections and reinfections, or (iii) the immunity induced by primary infections and reinfections. Point (i) means that we ignore both the infection-induced and vaccine-induced reduction in host infectiousness. Point (ii) means that we assume that infection-induced immunity does not shorten future infections, matching the analogous assumption for vaccine-induced immunity. This assumption is likely simplifying reality: hosts clear SARS-CoV-2 faster during reinfections [21]. While the exact implications for infectivity are unclear, prior immunity probably affects the duration of infectivity. Point (iii) implies that we ignore immune imprinting and, more generally, the accumulation of immunity from multiple infections. Hence, at the beginning of each reinfection, the immune status of each host is the same, irrespective of whether it is the first or a later reinfection.

Moreover, we do not account for hybrid immunity [6]. In the model with leaky immunity, the susceptibility to reinfection is independent of the host’s vaccination status. In other words, the vaccine-induced immunity does not protect recovered hosts, only the infection-induced immunity does. For the model with all-or-none immunity, there is a similar underlying assumption. If a host is infected, the probability of acquiring (full) protection against reinfection does not depend on their vaccination status. This assumption is also needed to obtain an analytical final-size expression (if vaccinees had a lower probability of becoming susceptible to reinfection, (5) would not hold). Neglecting hybrid immunity means that the results here underestimate the effects of vaccination in reducing the number of infections.

Finally, both models allow reinfections immediately after recovery from infection. There is some evidence suggesting that hosts may experience two infections in short succession with the same strain of Influenza A [26] and SARS-CoV-2 [34]. However, most reinfections likely occur due to a combination of antigenic escape [19, 35] and waning immunity [5]. We do not model waning immunity explicitly because we intended to focus on the dynamics of reinfection. Similarly, sequential reinfections (i.e., without waning of immunity) of the same strain are also possible in other models studying the evolution of Influenza A [12, 16] and SARS-CoV-2 [31, 33].

In this paper we have obtained final-size solutions of SIRI-type epidemic models with partial infection-induced immunity, which leads to reinfections. These models also include partial vaccine-induced protection against (primary) infections for a fraction of the population that has been vaccinated before the outbreak. Assuming all-or-none immunity, we obtain analytic final-size solutions in terms of the Lambert W function. These solutions have a similar mathematical structure to the final-size solution of the standard SIR model, but account for partial infection-induced immunity. Assuming instead leaky immunity, we find implicit solutions for the final epidemic size. These solutions (for a transient epidemic) are only valid if the susceptibility to reinfection is relatively low. If instead the susceptibility to reinfection is above a reinfection threshold, the system reaches an endemic state (similar to an SIS model). These results could help predict the epidemiological impact of disease that only induce partial protection in recovered hosts. For example, our final-size solutions quantify the reduction in the cumulative infections that would occur if a fraction of the population was vaccinated (prior to the outbreak). This reduction varies depending on the intrinsic transmissibility of the pathogen and the relative susceptibilities of vaccinees and recovered hosts.

## Data Availability

There is no data referred to in the manuscript.

## Statements and Declarations

We declare we have no competing interests. There are no supporting data.

## Acknowledgements

MAG was supported by the Gates Cambridge Trust (grant OPP1144 from the Bill & Melinda Gates Foundation). We are grateful for the comments of the anonymous reviewers. We especially appreciate that one of the reviewers provided us with alternative derivations of the final-size equations of this paper, which we will share to those who request it via email.

---

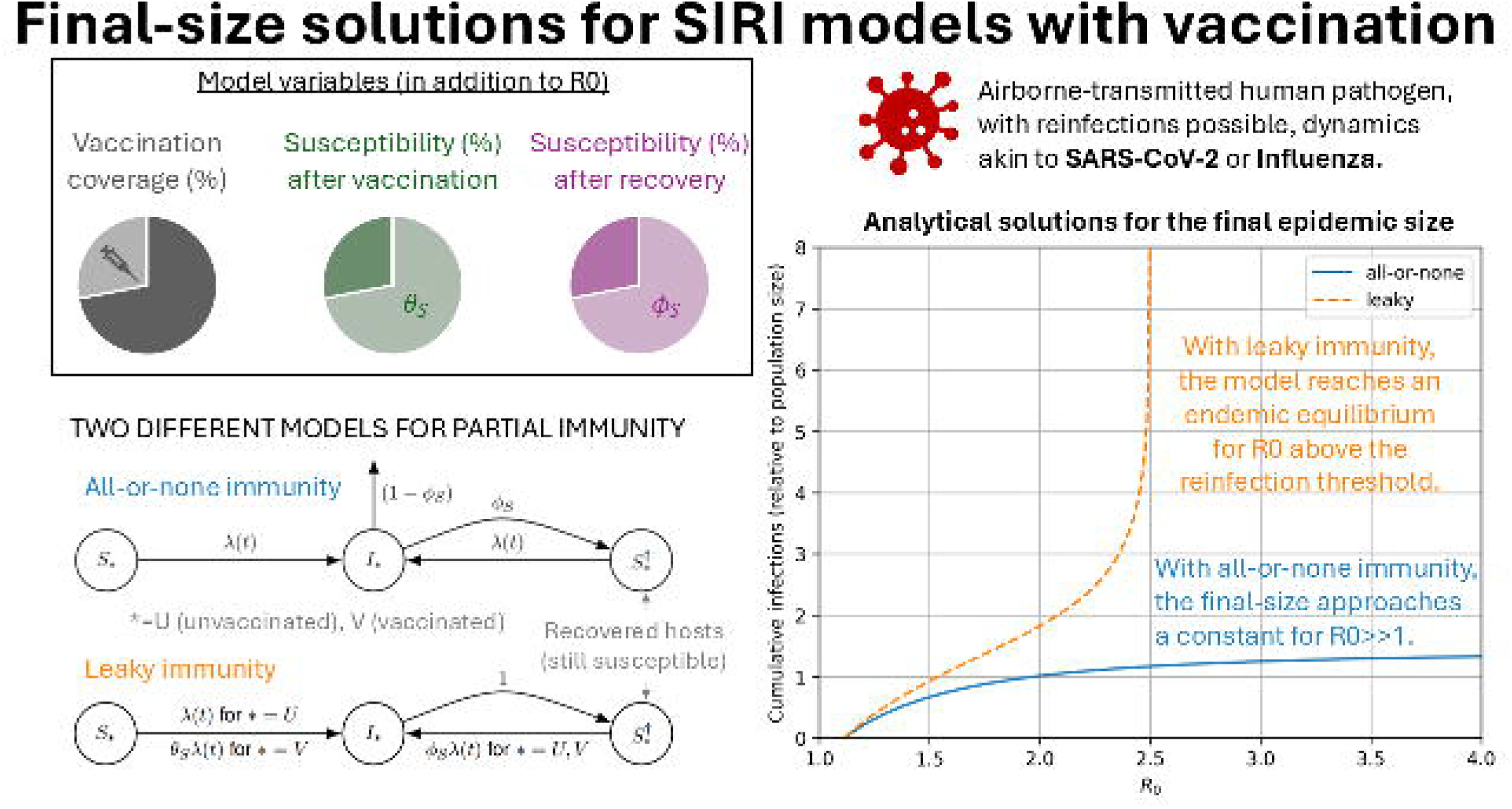

